# Links between cognition and functioning: Examining the role of mental health in clinically ascertained and population-based samples

**DOI:** 10.1101/2025.11.17.25340399

**Authors:** Amy J Lynham, Kimberley M Kendall, James TR Walters, Ian R Jones

**Affiliations:** National Centre for Mental Health, Division of Psychological Medicine and Clinical Neurosciences, School of Medicine, Cardiff University, Cardiff, United Kingdom

## Abstract

**Background:** Cognitive function is a significant predictor of health and mortality in the population. Common mental health problems, such as depression and anxiety, are associated with both cognitive impairments and increased functional impairment. This study aimed to examine the relationship between cognition, mental health and functioning across two cohorts.

**Methods:** Participants were recruited from an online population cohort, HealthWise Wales (N=3,679), and a psychiatric cohort, the National Centre for Mental Health (NCMH, N=1,036), to complete a cognitive battery and the World Health Organisation Disability Assessment Schedule (WHODAS). We assessed the associations between cognitive performance and the WHODAS, as well as two measures of life functioning: current employment and cohabitation with a partner. We examined the role of current mood, education and health/lifestyle factors using linear and logistic regression.

**Results:** Higher cognitive performance was associated with fewer functional difficulties on the WHODAS (HealthWise Wales: B=-0.12, 95%CI[-0.15,-0.09], P<0.001; NCMH: B=-0.1, 95%CI[-0.13,-0.08], P<0.001), being in employment or education (HealthWise Wales: OR=1.22, 95%CI[1.11,1.34], P<0.001; NCMH: OR=1.31, 95%CI[1.19,1.46], P<0.001) and living with a partner (HealthWise Wales only: OR=1.19, 95%CI[1.06,1.32], P=0.003). Higher levels of depression and anxiety symptoms were consistently associated with reduced functioning across all analyses. The relationship between cognition and functioning remained significant but attenuated after accounting for these symptoms.

**Conclusions:** Cognitive function was associated with functioning in both samples. This association may be partially explained by current symptoms of depression and anxiety. Both cognitive function and common mental health problems may be potential intervention targets to alleviate daily difficulties.

## Introduction

In 2021, an estimated 10 to 16 million people in the UK (18–24% of the population) were living with a disability, defined as the reduced ability to carry out daily activities for more than 12 months due to poor mental or physical health (1, 2). Mental health problems were the most reported cause of functional impairments among working adults with disabilities in 2021 (2). Individuals with disabilities in the UK face significant inequalities, including lower levels of education and employment, higher rates of poverty, loneliness, anxiety, and reduced life satisfaction (3). These disparities highlight the importance of understanding factors that contribute to functional impairments. Cognitive function may be one such determinant of daily functioning. Evidence from older adults shows that reduced cognitive ability is linked to difficulties with everyday activities such as mobility and self-care, while early-life cognitive function predicts long-term health outcomes (4–7). Although most research has focused on older populations, some studies have shown that cognitive function in young and middle-aged adults also predicts daily functioning, suggesting that the impact of cognition on functioning spans the entire adult lifespan (8–10).

Cognitive impairments are also closely linked to mental health disorders, which themselves are major contributors to disability, especially in working-age adults (11). Cognitive deficits have been identified in people with schizophrenia (12–14), bipolar disorder (15), depression (16), anxiety (17), and post-traumatic stress disorder (18), often affecting individuals’ ability to carry out life activities, maintain self-care, and participate in society (19–21). While studies in older adults have found connections between depressive symptoms, cognition, and daily functioning (22–25), less research has explored how mental health influences the cognition-functioning relationship in younger populations. Evidence from the Whitehall II and SWAN studies suggests that depressive symptoms have a modest attenuating effect on this relationship in middle-aged adults, but they did not examine anxiety or lifetime psychiatric diagnoses (8, 9). Given that mental disorders are a leading cause of disability globally (26), understanding their interaction with cognitive function is essential for addressing functional impairment across the adult lifespan.

### Current study

The aim of this cross-sectional study was to examine the relationship between cognition, mental health and life functioning in two Welsh cohorts: 1) HealthWise Wales, an online population sample, and 2) National Centre for Mental Health, a clinically ascertained psychiatric sample. We will use the broad terminology of “functioning” or “functional impairment” in recognition of the fact that not all social, occupational, or functional difficulties constitute a disability. We hypothesised that lower cognitive performance would be associated with greater functional difficulties, and current symptoms of depression and anxiety and psychiatric history would partly explain this.

## Methods

### Participants

#### National Centre for Mental Health

The National Centre for Mental Health (NCMH) is a Welsh Government-funded Research Centre that investigates mental health across the lifespan. Participants were invited to complete the study through letters or emails between October 2016 and July 2018 (N=6,904).

#### HealthWise Wales

HealthWise Wales is an online national population cohort of adults living in Wales or accessing their healthcare in Wales (27). The study was added as a module on the HealthWise Wales website in January 2020, and invitation emails were sent to all participants (N=29,492). Participants completed the study between January and December 2020.

All participants provided informed consent by checking boxes to indicate they agree to the consent statements online.

### Measures

#### Data Sources

In NCMH, all data were collected at a single time point, supplemented by existing diagnostic data from the wider NCMH project. In HealthWise Wales, data on cognition, self-reported functioning and symptoms of depression and anxiety were collected at a single time point. This was linked to existing HealthWise Wales data and the Secure Anonymised Information Linkage (SAIL) databank via the Secure Access Portal and Protected HWW Information Repository (SAPPHIRe), powered by the UK Secure e-Research Platform (UKSeRP) (28). SAIL is a national repository of anonymised health and administrative data for over 3 million people in Wales (29–33). Study data were imported into SAPPHIRe under existing permissions, adhering to research governance and data protection standards. HealthWise Wales data had been previously linked to SAIL (34), allowing us to use existing participant identifiers to match the study data without requiring identifiable information (3151 participants linked). Linked datasets included the Patient Episode Dataset for Wales (PEDW), the Welsh Longitudinal GP Dataset (WLGP), the Outpatient Database for Wales (OPDW), and the Emergency Department Dataset (EDDS). To mitigate the effects of using data collected at different timepoints, only HealthWise Wales questionnaires completed within one year prior to or following completion of the cognitive and functioning assessments were included.

#### Cognition

The Cardiff Online Cognitive Assessment (CONCA) was developed for psychiatric research (35). All tasks, including source codes, were developed by The Many Brains Project (36). NCMH participants completed CONCA between 2017 and 2019, during which time CONCA consisted of nine tasks hosted on The Many Brains Project’s TestMyBrain platform (35). CONCA was subsequently shortened to five tasks, (three core tasks and a further two optional tasks), and hosted on a custom-built website (37, 38). The tasks completed by each cohort are shown in Table 1.

**Table 1.** Cardiff ONline Cognitive Assessment (CONCA) tasks.

| **Task** | **Domain** | **NCMH** | **HealthWise Wales** |
| --- | --- | --- | --- |
| **Digit Symbol Coding** | Processing Speed | X | X |
| **Backward Digit Span** | Working Memory | X | X |
| **Vocabulary** | Crystallised IQ | X | X |
| **Morphed Emotion Identification** | Social Cognition | X | X |
| **Matrix Reasoning** | Executive Function | X | X |
| **Verbal Paired Associates** | Verbal Learning | X |  |
| **Hartshorne Visual Working Memory** | Visual Learning | X |  |
| **Balloon Analogue Risk Task** | Strategic Risk Taking | X |  |
| **Multiple Object Tracking** | Attention | X |  |

NCMH completed nine CONCA tasks (2017-2019). HealthWise Wales completed five CONCA tasks in 2020, two of which were optional (Morphed Emotion Identification and Matrix Reasoning).

#### Functioning

Daily functional impairments were assessed using the World Health Organisation Disability Assessment Schedule, Version 2 (WHODAS 2.0 (39)). WHODAS 2.0 measures difficulties over the past 30 days across six domains: mobility, self-care, household responsibilities, subjective cognition, social interactions and participation in society, with responses rated from “none” to “extreme or cannot do”. It should be noted that the WHODAS 2.0 was an optional questionnaire for HealthWise Wales, which was completed by 1033/3679 participants (28%). A comparison of HealthWise Wales participants who completed the WHODAS and those who did not is provided in Supplementary Table S1. Two additional binary variables were included: (1) living with a partner and (2) employed or in education (for participants under 66 years).

#### Mental health and lifestyle factors

Mental health and lifestyle variables were selected based on existing literature for their association with cognition and functioning: current depression and anxiety symptoms (26, 40, 41), history of psychiatric diagnoses (42), educational attainment (10, 43), body mass index (BMI) (10, 44, 45), alcohol use (10, 46), ever smoked (10, 47, 48), and physical activity (49, 50). We attempted to select variables from both samples that were as closely matched as possible. Measures and sources of data for each variable are outlined in Table 2.

**Table 2.** Measures across studies.

|  | NCMH |  | HealthWise Wales |  |
| --- | --- | --- | --- | --- |
|  | Measure | Source | Measure | Source |
| **Self-reported functioning** | WHODAS | CONCA<sup>1</sup> | WHODAS | CONCA (optional) |
| **Employment** | Currently employed or in education (yes/no) | CONCA | Currently employed or in education (yes/no) | HealthWise Wales |
| **Cohabiting** | Not collected |  | Currently living with a partner (yes/no) | HealthWise Wales |
| **History of psychiatric diagnoses** | Self-report of 1) psychosis, 2) bipolar disorder, 3) depression, 4) anxiety, 5) OCD, 6) PTSD, 7) eating disorder, 8) personality disorder, 9) autism, 10) ADHD or 11) other mental health problems (yes/no) | CONCA and NCMH | History of depression or anxiety (yes/no) | SAIL Databank |
| **Current depression and anxiety symptoms** | Hospital Anxiety and Depression Scale (51) (total score) | CONCA | Hospital Anxiety and Depression Scale (51) (total score) | CONCA (optional) |
| **Education** | University degree (yes/no) | CONCA | University degree (yes/no) | CONCA |
| **History of smoking** | Ever smoked (yes/no) | CONCA (corroborated against NCMH) | Ever smoked (yes/no) | HealthWise Wales |
| **Alcohol use** | 1) Current regular alcohol use (yes/no) | CONCA | Evidence of binge drinking based on UK Government guidelines (yes/no) | HealthWise Wales |
|  | 2) self-report of alcohol or substance use disorder | CONCA and NCMH |  |  |
| **Body Mass Index** | Insufficient data, as BMI was not included in all versions of NCMH's questionnaire |  | BMI | HealthWise Wales |
| **Physical Activity** | Not collected |  | General Practice Physical Activity Questionnaire (52) (active, moderately active, moderately inactive or inactive)<sup>2</sup> | HealthWise Wales |

<sup>1</sup>CONCA refers to the current study and indicates that the data was collected concurrently with the cognitive assessment; <sup>2</sup>Physical activity includes planned physical exercise (e.g. gym, sports), cycling, walking, housework, gardening and DIY, as well as the type and amount of physical activity involved in their work. CONCA, Cardiff ONline Cognitive Assessment; NCMH, National Centre for Mental Health; SAIL, Secure Anonymised Information Linkage; WHODAS, World Health Organisation Disability Assessment Schedule

### Psychiatric history

In NCMH, psychiatric diagnoses were primarily self-reported in response to the question, “Has a doctor or health professional ever told you that you have any of the following diagnoses?” Participants selected all diagnoses that applied. Additional data were obtained from other NCMH projects, which used self-report and semi-structured interviews (see Supplementary Note 1). Consistent with previous NCMH studies (53, 54), the following diagnoses were included as binary variables: psychosis, bipolar disorder, depression, anxiety, obsessive-compulsive disorder, post-traumatic stress disorder, eating disorder, personality disorder, alcohol or substance use disorder, autism spectrum disorder, attention deficit hyperactivity disorder and other mental health problems. Diagnoses were not mutually exclusive so participants could be included in multiple groups. This was not the case for individuals with psychosis or bipolar disorder, who were excluded from the depression and anxiety groups.

In HealthWise Wales, we derived data on psychiatric diagnoses from electronic health records held in the SAIL databank. Participants were rated as having a diagnosis of depression or anxiety if they had a historical diagnosis of depression/anxiety *and* there was a record of treatment *and* a record of diagnosis or symptoms in their medical records within 12 months of completing CONCA using the algorithm described by John et al. (55) (algorithm 10).

### Analysis

All analyses were conducted using R version 3.6.1 (56). Z scores were derived for each cognitive task, and general cognitive ability (‘g’) was derived using multidimensional scaling (an analogous approach to principal component analysis that can accommodate missing data), including scores on all the tasks (57). WHODAS domains were derived through confirmatory factor analysis using the ‘lavaan’ package, based on theoretical models for the 12-item WHODAS (58, 59), as the scoring procedure for the 12-item version produces an overall score only. Diagonally weighted least squares estimation was used, as this is recommended for ordinal data (60). Model fit was evaluated using criteria based on previous research (CFI>0.9, TLI>0.9, RMSEA<0.08, SRMR<0.08) (58, 59), and was improved by allowing covariance between latent factors based on modification indices. Factor scores were derived from the most appropriate model.

Linear and logistic regressions were conducted to assess the associations between cognitive performance and 1) WHODAS factor scores, 2) living with a partner or spouse, and 3) employed or in full-time education. Each cognitive task and ‘g’ were analysed separately, with age and gender as covariates; device type was also included for the HealthWise Wales sample (not available for NCMH). P values were corrected using the false discovery rate (FDR) method.

To assess potential confounding mental health and lifestyle factors, we examined the associations between each variable and 1) cognitive performance and 2) functioning using linear or logistic regression (see Supplementary Table 1). Variables significantly associated with both were added as covariates in follow-up models. Separate models were run for each outcome (WHODAS, employment, cohabiting) in each sample, including all relevant confounding variables.

## Results

### Sample characteristics

The total number of participants who completed at least one cognitive task was 1,036 in NCMH (16-79 years) and 3,679 in HealthWise Wales (16-93 years). Completion rates varied across the variables (see Table 3). Compared to the HealthWise Wales sample, the NCMH participants were significantly younger (t(4702)=18.17, P<0.001), had higher WHODAS scores (t(2001)=-15.11, P<0.001), higher HADS scores (t(2068)=-15.05, P<0.001), lower rates of employment (X^2^=66.96, P<0.001), fewer undergraduate degrees (X^2^=7.95, P=0.005) and were less likely to report that they had ever smoked (X^2^=41.77, P<0.001). The NCMH participants had lower performance than the HealthWise Wales participants on Digit Symbol Coding (t(4629)=8.04, P<0.001), Vocabulary (t(3854)=14.79, P<0.001), Matrix Reasoning (t(3190)=5.17, P<0.001) and higher performance on Backward Digit Span (t(3999)=-4.27, P<0.001). The samples did not differ in gender distributions (X^2^=1.04, P=0.31) or scores on the Morphed Emotion Identification task (t(3171)=-0.6, P=0.55).

**Table 3.**
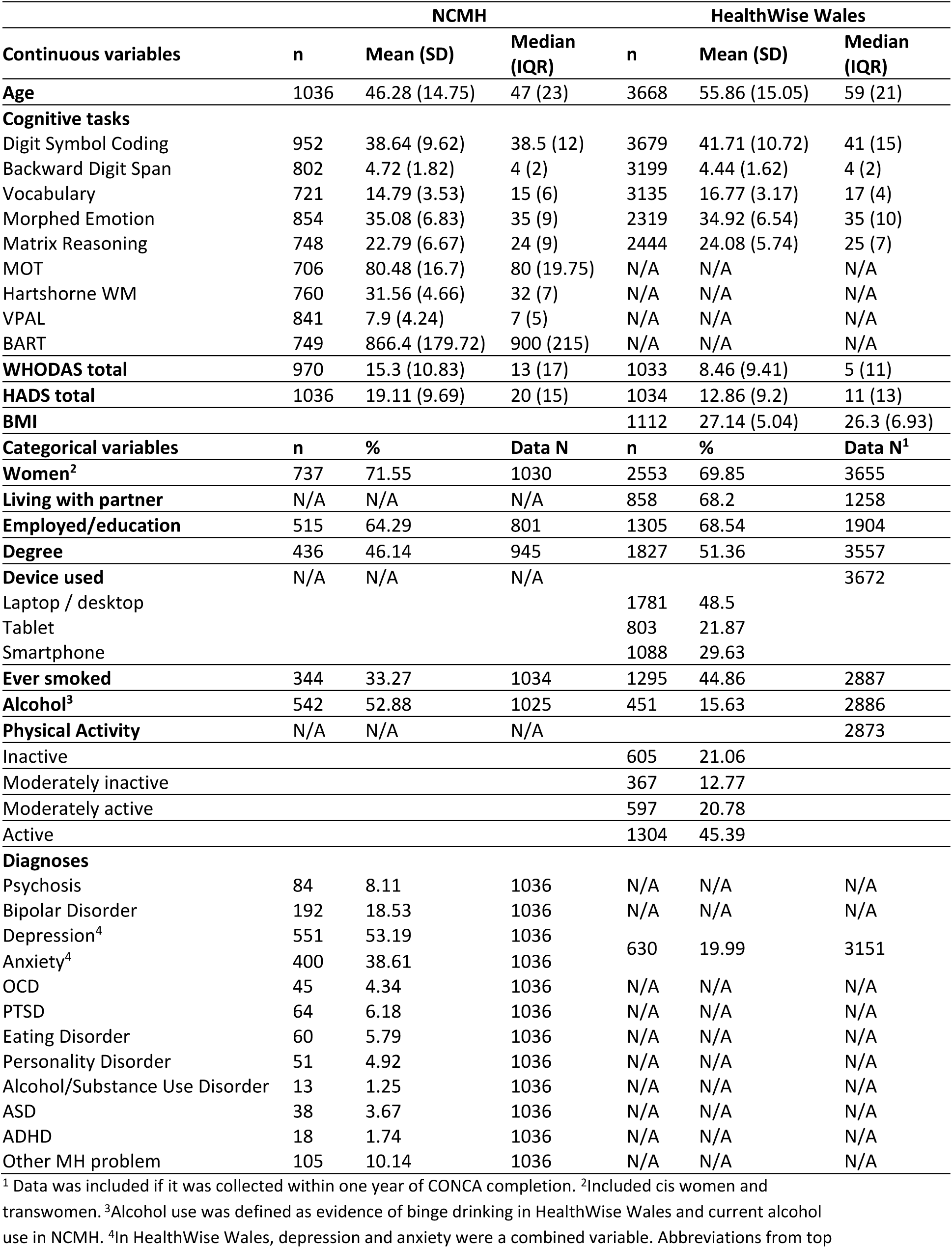
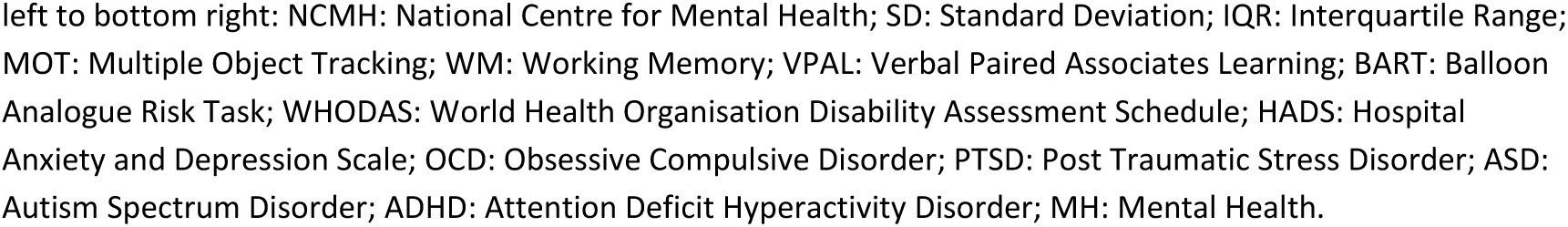
Sample characteristics.

| Continuous variables | NCMH |  |  | HealthWise Wales |  |  |
| --- | --- | --- | --- | --- | --- | --- |
|  | n | Mean (SD) | Median (IQR) | n | Mean (SD) | Median (IQR) |
| **Age** | 1036 | 46.28 (14.75) | 47 (23) | 3668 | 55.86 (15.05) | 59 (21) |
| **Cognitive tasks** |  |  |  |  |  |  |
| Digit Symbol Coding | 952 | 38.64 (9.62) | 38.5 (12) | 3679 | 41.71 (10.72) | 41 (15) |
| Backward Digit Span | 802 | 4.72 (1.82) | 4 (2) | 3199 | 4.44 (1.62) | 4 (2) |
| Vocabulary | 721 | 14.79 (3.53) | 15 (6) | 3135 | 16.77 (3.17) | 17 (4) |
| Morphed Emotion | 854 | 35.08 (6.83) | 35 (9) | 2319 | 34.92 (6.54) | 35 (10) |
| Matrix Reasoning | 748 | 22.79 (6.67) | 24 (9) | 2444 | 24.08 (5.74) | 25 (7) |
| MOT | 706 | 80.48 (16.7) | 80 (19.75) | N/A | N/A | N/A |
| Hartshorne WM | 760 | 31.56 (4.66) | 32 (7) | N/A | N/A | N/A |
| VPAL | 841 | 7.9 (4.24) | 7 (5) | N/A | N/A | N/A |
| BART | 749 | 866.4 (179.72) | 900 (215) | N/A | N/A | N/A |
| **WHODAS total** | 970 | 15.3 (10.83) | 13 (17) | 1033 | 8.46 (9.41) | 5 (11) |
| **HADS total** | 1036 | 19.11 (9.69) | 20 (15) | 1034 | 12.86 (9.2) | 11 (13) |
| **BMI** |  |  |  | 1112 | 27.14 (5.04) | 26.3 (6.93) |
| **Categorical variables** | **n** | **%** | **Data N** | **n** | **%** | **Data N<sup>1</sup>** |
| **Women<sup>2</sup>** | 737 | 71.55 | 1030 | 2553 | 69.85 | 3655 |
| **Living with partner** | N/A | N/A | N/A | 858 | 68.2 | 1258 |
| **Employed/education** | 515 | 64.29 | 801 | 1305 | 68.54 | 1904 |
| **Degree** | 436 | 46.14 | 945 | 1827 | 51.36 | 3557 |
| **Device used** | N/A | N/A | N/A |  |  | 3672 |
| Laptop / desktop |  |  |  | 1781 | 48.5 |  |
| Tablet |  |  |  | 803 | 21.87 |  |
| Smartphone |  |  |  | 1088 | 29.63 |  |
| **Ever smoked** | 344 | 33.27 | 1034 | 1295 | 44.86 | 2887 |
| **Alcohol<sup>3</sup>** | 542 | 52.88 | 1025 | 451 | 15.63 | 2886 |
| **Physical Activity** | N/A | N/A | N/A |  |  | 2873 |
| Inactive |  |  |  | 605 | 21.06 |  |
| Moderately inactive |  |  |  | 367 | 12.77 |  |
| Moderately active |  |  |  | 597 | 20.78 |  |
| Active |  |  |  | 1304 | 45.39 |  |
| **Diagnoses** |  |  |  |  |  |  |
| Psychosis | 84 | 8.11 | 1036 | N/A | N/A | N/A |
| Bipolar Disorder | 192 | 18.53 | 1036 | N/A | N/A | N/A |
| Depression<sup>4</sup> | 551 | 53.19 | 1036 | 630 | 19.99 | 3151 |
| Anxiety<sup>4</sup> | 400 | 38.61 | 1036 |  |  |  |
| OCD | 45 | 4.34 | 1036 | N/A | N/A | N/A |
| PTSD | 64 | 6.18 | 1036 | N/A | N/A | N/A |
| Eating Disorder | 60 | 5.79 | 1036 | N/A | N/A | N/A |
| Personality Disorder | 51 | 4.92 | 1036 | N/A | N/A | N/A |
| Alcohol/Substance Use Disorder | 13 | 1.25 | 1036 | N/A | N/A | N/A |
| ASD | 38 | 3.67 | 1036 | N/A | N/A | N/A |
| ADHD | 18 | 1.74 | 1036 | N/A | N/A | N/A |
| Other MH problem | 105 | 10.14 | 1036 | N/A | N/A | N/A |

<sup>1</sup> Data was included if it was collected within one year of CONCA completion. <sup>2</sup>Included cis women and transwomen. <sup>3</sup>Alcohol use was defined as evidence of binge drinking in HealthWise Wales and current alcohol use in NCMH. <sup>4</sup>In HealthWise Wales, depression and anxiety were a combined variable. Abbreviations from top
left to bottom right: NCMH: National Centre for Mental Health; SD: Standard Deviation; IQR: Interquartile Range; MOT: Multiple Object Tracking; WM: Working Memory; VPAL: Verbal Paired Associates Learning; BART: Balloon Analogue Risk Task; WHODAS: World Health Organisation Disability Assessment Schedule; HADS: Hospital Anxiety and Depression Scale; OCD: Obsessive Compulsive Disorder; PTSD: Post Traumatic Stress Disorder; ASD: Autism Spectrum Disorder; ADHD: Attention Deficit Hyperactivity Disorder; MH: Mental Health.

In HealthWise Wales, participants who completed the WHODAS (n=1,033) were younger (B=-6.82, 95%CI[-7.88,-5.76], P<0.001), more likely to be women (X^2^=13.46, P<0.001), more likely to have a degree (X^2^=14.1, P<0.001), less likely to smoke (X^2^=6.01, P=0.01), less likely to use a tablet computer (X^2^=9.37, P=0.009) and more likely to have a history of depression or anxiety (X^2^=6.43, P=0.01) than participants who did not complete the WHODAS (see Supplementary Table S2). After correction for age, gender and device, participants who completed the WHODAS had higher scores on Digit Symbol Coding than participants who did not (B=0.08, 95%CI[0.03,0.14], P=0.01), but there were no other significant differences in cognitive scores.

### WHODAS

#### Confirmatory factor analysis

In both samples, the best-fitting WHODAS model included an overall functional impairment factor and six domains: mobility, self-care, household responsibilities, cognition, social, and society (see Supplementary Table S3). Model fit was improved by allowing covariation between related latent factors, primarily those linked to physical mobility, which was guided by modification indices and similar models in previous research (59). The final model for both samples yielded the following fit indices: CFI=1, TLI=1, RMSEA<0.05, SRMR<0.05 (see Supplementary Figure 1). Factor scores were derived for overall functioning and each domain.

#### Associations with cognitive performance

Higher general cognitive ability (‘g’) was associated with better overall functioning in NCMH (B=-0.1, 95%CI[-0.13,-0.08], P<0.001) and HealthWise Wales (B=-0.12, 95%CI[-0.15,-0.09], P<0.001) across all WHODAS domains (see Table 4). The associations between ‘g’ and overall functioning remained significant when the WHODAS cognition domain was removed (NCMH: B=-0.09, 95%CI[-0.11,-0.07], P<0.001; HealthWise Wales: B=-0.12, 95%CI[-0.15,-0.09], P<0.001). Associations between the individual tasks and overall functioning are available in Supplementary Table S4. Due to WHODAS scores skewed towards 0 in the HealthWise Wales sample, we conducted follow-up analyses using dichotomised domains (any impairment vs. none). Higher ‘g’ was associated with lower likelihood of impairment across all domains (OR=0.65-0.83, see Supplementary Table S5). Finally, we repeated the analyses restricting the sample to participants of working age (<66 years, NCMH n=868, HealthWise Wales n=801). This did not change the results (see Table 4 and Supplementary Table S4).

**Table 4.** Associations between functioning domains and ’g’ in HealthWise Wales and NCMH.

| Domain | HealthWise Wales |  |  | National Centre for Mental Health |  |  |
| --- | --- | --- | --- | --- | --- | --- |
|  | B | 95% CI | P | B | 95% CI | P |
| **All Participants** |  |  |  |  |  |  |
| Overall impairment | -0.12 | -0.15, -0.09 | <0.001 | -0.1 | -0.13, -0.08 | <0.001 |
| Mobility | -0.15 | -0.2, -0.1 | <0.001 | -0.14 | -0.18, -0.1 | <0.001 |
| Self-care | -0.13 | -0.17, -0.09 | <0.001 | -0.13 | -0.16, -0.09 | <0.001 |
| Household | -0.15 | -0.19, -0.11 | <0.001 | -0.14 | -0.18, -0.11 | <0.001 |
| Cognition | -0.12 | -0.15, -0.09 | <0.001 | -0.15 | -0.18, -0.11 | <0.001 |
| Social | -0.15 | -0.19, -0.11 | <0.001 | -0.18 | -0.22, -0.14 | <0.001 |
| Society | -0.17 | -0.21, -0.13 | <0.001 | -0.19 | -0.23, -0.14 | <0.001 |
| **Participants aged 65 years and younger** |  |  |  |  |  |  |
| Overall impairment | -0.13 | -0.17, -0.1 | <0.001 | -0.1 | -0.12, -0.08 | <0.001 |
| Mobility | -0.16 | -0.21, -0.11 | <0.001 | -0.13 | -0.17, -0.09 | <0.001 |
| Self-care | -0.14 | -0.19, -0.1 | <0.001 | -0.12 | -0.16, -0.09 | <0.001 |
| Household | -0.17 | -0.21, -0.12 | <0.001 | -0.14 | -0.17, -0.1 | <0.001 |
| Cognition | -0.14 | -0.17, -0.1 | <0.001 | -0.14 | -0.17, -0.11 | <0.001 |
| Social | -0.17 | -0.22, -0.12 | <0.001 | -0.18 | -0.22, -0.13 | <0.001 |
| Society | -0.19 | -0.24, -0.14 | <0.001 | -0.18 | -0.23, -0.14 | <0.001 |

Results from linear regressions with WHODAS domains and overall functional impairment as outcome measures.
B: regression coefficient; 95%CI: 95% confidence intervals; P: adjusted p-value.

#### Role of mental health and other covariates

In NCMH, the variables associated with both ‘g’ and WHODAS were HADS scores, current alcohol use, ever smoked, education and diagnosis of bipolar disorder (see Supplementary Table S1). After inclusion of these covariates, higher ‘g’ remained significantly associated with higher overall functioning (B=-0.03, 95%CI[-0.05,-0.02], P<0.001). Higher HADS score (B=0.04, 95%CI[0.04,0.05], P<0.001) and diagnosis of bipolar disorder (B=0.13, 95%CI[0.06,0.2], P<0.001) were associated with lower overall functioning.

In HealthWise Wales, the variables associated with both ’g’ and WHODAS were HADS scores, BMI, ever smoked, physical activity, education and diagnosis of anxiety/depression (see Supplementary Table S1). After inclusion of these covariates, higher ‘g’ remained significantly associated with higher overall functioning (B=-0.05, 95%CI[-0.08,-0.01], P=0.02). Higher HADS score (B=0.05, 95%CI[0.04,0.05], P<0.001) and history of anxiety/depression (B=0.16, 95%CI[0.04,0.29], P=0.009) were associated with lower overall functioning, whereas being active (B=-0.14, 95%CI[-0.26,-0.01], P=0.03) was associated with higher functioning. These results were consistent across the domains of functioning in both samples (see Supplementary Table S6), with the exception that in HealthWise Wales, cognition was no longer associated with the WHODAS social domain after inclusion of covariates.

### Real-world indicators of functioning

Higher general cognitive ability (‘g’) was associated with being in employment or education in NCMH (OR=1.31, 95%CI[1.19,1.46], P<0.001). Data on living with a partner were not available in NCMH. Higher ‘g’ was associated with a higher likelihood of living with a partner or spouse (OR=1.19, 95%CI[1.06,1.32], P=0.003) and being in employment or education (OR=1.22, 95%CI[1.11,1.34], P<0.001) in HealthWise Wales. Associations between individual tasks and outcome measures are available in Supplementary Table S4.

#### Role of mental health and other covariates

In NCMH, the variables associated with both ‘g’ and current employment were HADS scores, current alcohol use, ever smoked, education and diagnoses of bipolar disorder, psychosis or depression. After inclusion of these covariates, ‘g’ was not associated with employment (OR=1.09, 95%CI[0.97,1.23], P=0.15). Higher HADS score (OR=0.93, 95%CI[0.9,0.95], P<0.001), diagnosis of bipolar disorder (OR=0.52, 95%CI[0.27,0.99], P=0.047) and diagnosis of psychosis (OR=0.32, 95%CI[0.14,0.73], P=0.007) were associated with lower likelihood of employment. Current alcohol use (OR=1.5, 95%CI[1,2.24], P=0.048) and having a degree (OR=2.03, 95%CI[1.34,3.06], P<0.001) were associated with higher likelihood of employment.

In HealthWise Wales, the variables associated with both ‘g’ and current employment were HADS scores, BMI, ever smoked, physical activity, education and diagnosis of anxiety/depression (see Supplementary Table S1). After inclusion of these covariates, ‘g’ was not significantly associated with employment (OR=1, 95%CI[0.73,1.38], P=1). Higher HADS scores were associated with a reduced likelihood of employment (OR=0.94, 95%CI[0.89,0.98], P=0.007). Being moderately inactive (OR=5.9, 95%CI[1.64,24.56], P=0.009) and active (OR=3.7, 95%CI[1.41,10], P=0.009) were associated with an increased likelihood of employment compared to being inactive. The variables associated with both ‘g’ and cohabiting were HADS scores, ever smoked, physical activity and education. After inclusion of these covariates, ‘g’ was significantly associated with cohabitating (OR=1.15, 95%CI[1.03,1.3], P=0.02). History of smoking was associated with a reduced likelihood of cohabiting (OR=0.73, 95%CI[0.55,0.96], P=0.02).

## Discussion

This cross-sectional study examined the relationships between cognition, mental health, and functioning. Lower cognitive scores were associated with more self-reported difficulties and reduced likelihood of cohabitation and employment. Depression and anxiety symptoms were consistently associated with cognition and functioning across clinical and population samples.

### WHODAS

Lower cognitive performance was associated with greater difficulties in participants’ mobility, household management, self-care, self-reported cognitive function, participation in their community and social relationships over the previous 30 days. These findings align with previous research showing that cognition is associated with daily functioning, including personal care, lifting and carrying, walking, managing a household, managing medical care, and getting around (8–10). This study extends prior research by including younger adults, as 38% of NCMH and 31% of HealthWise Wales participants were aged 40 or under. The association between cognition and functioning remained significant after controlling for age and when limited to working-age adults, suggesting the findings are not driven solely by age-related decline. Whilst Jacob et al. (10) included participants as young as 16 years, they relied on the National Adult Reading Test (NART), a measure of crystallised IQ. Our study employed a broader cognitive battery that incorporated both crystallised and fluid intelligence. Fluid intelligence may be a stronger predictor of health outcomes, including mortality (61). Indeed, the associations between vocabulary (crystallised intelligence) and the WHODAS were among the weakest across the two samples (though still significant).

Another strength of this study was the use of the same cognitive and outcome measures in both a clinical sample with diagnosed mental health conditions and a population-based sample. In both datasets, a one standard deviation (1SD) increase in overall cognition ‘g’ was associated with decreases of 17-35% in the odds of reporting impairment across the six domains. The NCMH is a sample of participants with diagnosed psychiatric disorders, including psychotic, mood and anxiety disorders, so cognitive and functional impairments are more likely and thus, our results are not unexpected. In contrast, the participants recruited from HealthWise Wales were an online population sample with high levels of education and employment (a full comparison of this sample compared with the Welsh population estimates can be found in Lynham et al. (38)). Despite this, the strengths of these associations were similar across both the clinical and population-derived datasets.

As a cross-sectional study, we cannot determine the direction of causality between cognition and functioning. Functional impairments (e.g. limited motor function) could affect cognitive performance, and various health conditions are known to impact cognition (16, 62–64). Cognitive ability also influences how well individuals understand and manage their health conditions, as more complex health information increases cognitive burden, reducing the likelihood it is retained (65, 66). In chronic conditions, including diabetes, asthma, and COPD, processing speed and working memory have been associated with health literacy and subsequent health management (67–70). Thus, poorer cognition may lead to functional impairment due to inadequate health management. Longitudinal research has shown that cognitive function can predict later physical function, but not the reverse (8).

In both datasets, current depression and anxiety symptoms were strongly associated with WHODAS scores. This may partly reflect the commonalities between the HADS and WHODAS, as both assess recent self-reported issues with emotions, self-care and daily activities. Individuals with these symptoms may also be more likely to report difficulties due to negative bias. However, the similar effect of HADS on cognition and employment analysis suggests that the effect extends beyond reporting bias. Cognitive and functional impairments may result from mood and anxiety disorders (62), though it’s also possible that lower cognition increases vulnerability to mood symptoms and functional impairments.

In HealthWise Wales, both history of depression/anxiety *and* current symptoms were associated with poorer functioning. In NCMH, history of depression or anxiety was not included in the final model, as neither diagnosis was associated with WHODAS scores. This is likely because NCMH is a clinical sample, so those without depression or anxiety will have other (possibly more severe) mental health conditions. In contrast, HealthWise Wales is a population-based sample, so we were comparing participants with a diagnosis of depression/anxiety against predominantly those without a history of psychiatric disorders. As such, the analyses across the two samples were addressing slightly different questions. In NCMH, diagnosis of bipolar disorder was associated with both cognitive performance and WHODAS scores, which is consistent with studies showing poor cognitive and functional outcomes in bipolar disorder (19).

Lifestyle factors that may underly the association between cognitive function and self-reported functional impairment were also investigated in the current study, including alcohol use, smoking, BMI and physical activity (the latter two in HealthWise Wales only). In HealthWise Wales, physical activity was the only factor associated with better overall functioning. In NCMH, neither smoking nor alcohol use was significantly associated with functioning in the final model.

Finally, it is worth noting that the association between cognition and WHODAS scores remained significant after controlling for these covariates. This may suggest a direct association between cognition and functioning, or that other explanatory variables were not included in this study. This study focused on the role of mental health, but future studies should evaluate the role of physical health, particularly given evidence that some common physical health problems impact cognition (63, 64).

### Real-world indicators of functioning

Two real-world functioning indicators were associated with cognition: living with a partner or spouse and being in employment or education. In HealthWise Wales, an increase of 1SD in ‘g’ was associated with a 19% increase in the likelihood of cohabiting. Few studies have examined the relationship between cohabitation/marriage and current cognitive function; however, one study of healthy, middle-aged adults found that married individuals had better episodic memory than single individuals (71). The authors proposed that the social interactions involved in marriage may stimulate cognitive function by facilitating neuronal growth and maintenance, as well as protecting against degeneration (71).

Our finding that cognitive function is associated with employment status is consistent with previous research, which demonstrates that employed participants have higher cognitive scores than unemployed participants (72). An increase of 1SD in ‘g’ was associated with a 22% increase in the likelihood of employment in

HealthWise Wales and a 31% increase in NCMH. In both datasets, the association between cognition and employment was not significant after adjusting for covariates. In NCMH, higher HADS scores and diagnosis of bipolar disorder or psychosis were associated with a reduced likelihood of employment, whilst current alcohol use and having a degree were associated with higher likelihood of employment. Thus, the relationship between lower cognition and reduced employment appears to be explained by education and illness-related factors, such as current depressive and anxiety symptoms and diagnosis of severe mental illness. This also seemed to be the case in HealthWise Wales, where the association was not significant after inclusion of current depression/anxiety symptoms and physical activity.

### Limitations

This study has several limitations beyond its cross-sectional design. Online data collection can lead to recruitment bias, as we have previously demonstrated (35, 38). In the HealthWise Wales cohort, the WHODAS and HADS were optional, which limited the sample size and generalisability. The subsample who completed the WHODAS was younger, more likely to be women, and had a higher degree of education, and was less likely to smoke or use a tablet computer. This may mean they were less likely to report functional impairments than the wider cohort. Nonetheless, we obtained WHODAS data on over 1,000 participants, 22-67% of whom reported impairment across domains, and found an association between cognition and WHODAS scores.

Data from CONCA were integrated with existing HealthWise Wales data, resulting in some variables being collected at different times. To address this, we included only data collected within one year of CONCA participation. Importantly, variables such as BMI, alcohol use and physical activity are generally stable over time (73, 74).

While the HADS aims to distinguish between depression and anxiety, the high correlation between subscales and frequent co-occurrence of both disorders meant we could not isolate their effects. However, given the high comorbidity seen in the general population, focusing on these symptoms together may better reflect real-world experiences (75).

Both HealthWise Wales and NCMH recruited in the same regions simultaneously, so some sample overlap is possible. We were unable to determine the extent of this due to data-sharing restrictions. Any overlap is likely to be minimal, as this would require participation in four separate studies (HealthWise Wales, NCMH and two CONCA follow-ups).

### Conclusions

In summary, we report that cognitive performance was associated with measures of functioning in a population sample and a psychiatric sample. A measure of current depression and anxiety symptoms was identified as a key factor in this relationship, highlighting the important contribution of common mental health symptoms towards functioning. Functional impairments have personal, societal, and economic consequences; thus, identifying factors that might contribute is important. Both cognitive function and depression/anxiety symptoms may be valuable targets for interventions designed to improve functional outcomes, not just in populations of people with psychiatric disorders but more widely. Longitudinal approaches are needed to determine the causal direction of the associations between cognition and functioning.

## Statement of Ethical Approval

The authors assert that all procedures contributing to this work comply with the ethical standards of the relevant national and institutional committees on human experimentation and with the Helsinki Declaration of 1975, as revised in 2013. All procedures involving human participants were approved by: Cardiff University’s School of Medicine Research Ethics Committee (CONCA, reference: 15/64), Wales Research Ethics Committee 3 (HealthWise Wales, reference: 15/WA/0076) and Wales Research Ethics Committee 2 (NCMH, reference: 16/WA/0323).

## Data Availability

CONCA data on NCMH participants are available online in the MRC Centre for Neuropsychiatric Genetics and Genomics Walters Group Data Repository at https://walters.psycm.cf.ac.uk. Authors are not able to share data from the NCMH or HealthWise Wales cohorts directly due to restrictions. Data on the wider NCMH sample are available through direct application to NCMH (see https://www.ncmh.info for contact details). The HealthWise Wales dataset is available through HealthWise Wales on reasonable request (https://www.healthwisewales.gov.wales/for-researchers/).

## Acknowledgements

This study was facilitated by HealthWise Wales, the Health and Care Research Wales initiative, which is led by Cardiff University in collaboration with SAIL, Swansea University. Responsibility for the interpretation of the information provided in this manuscript is the authors’ alone. The authors thank Dr Pauline Ashfield-Watt for her support with recruitment and linkage for this study.

This research was conducted with the support of The Many Brains Project, Inc. The web-based tasks remain the intellectual property of The Many Brains Project, which retains the copyright for these tasks and the source code. The authors thank Dr Laura Germine for her support in developing the CONCA battery.

This study makes use of anonymised data held in the Secure Anonymised Information Linkage (SAIL) Databank. We would like to acknowledge all the data providers who make anonymised data available for research. The responsibility for the interpretation of the information supplied is the authors’ alone.

The authors thank our public involvement contributors who co-designed the CONCA battery and supported the study, including our CONCA advisory group, members of NCMH-PAR and the health professionals of Headroom, Cardiff & Vale University Health Board.

The authors also thank the HealthWise Wales and NCMH study participants for their invaluable contribution to this project.

## Author Contributions

AJL formulated the research question and designed the study with input from all co-authors. AJL selected and created the research assessments and collected the data, with input from IRJ and JTRW. AJL and KMK developed the analysis plan. AJL conducted the data analysis. AJL wrote the draft of the manuscript and incorporated revisions by co-authors. All authors reviewed the manuscript for intellectual content, contributed to revisions and approved the final version for publication.

## Transparency Declaration

As lead author, I affirm that the manuscript is an honest, accurate and transparent account of the study being reported; that no aspects of the study have been omitted and that any discrepancies from the study as planned have been explained.

## Data Availability

CONCA data on NCMH participants are available in the MRC Centre for Neuropsychiatric Genetics and Genomics Walters Group Data Repository, https://walters.psycm.cf.ac.uk. Authors are not able to share data from the NCMH or HealthWise Wales cohorts directly due to restrictions. Data on the wider NCMH sample are available through direct application to NCMH (see https://www.ncmh.info for contact details). The HealthWise Wales dataset is available through HealthWise Wales on reasonable request (https://www.healthwisewales.gov.wales/for-researchers/).

## Analytic Code Availability

The analytic code used to generate these findings of this study are available from the corresponding author, AJL, upon reasonable request, with the exception of the algorithms used to generate diagnoses from SAIL, which must be requested through SAIL (https://saildatabank.com/).

