## Supplementary for "Links between cognition and functioning: Examining the role of mental health in clinically ascertained and population-based samples"

**Running Title**: Cognition, functioning, and mental health

**Authors**:

Amy J Lynham, PhD, Kimberley M Kendall, PhD, James TR Walters, PhD, Ian R Jones, PhD

**Affiliations**:

Cardiff Centre for Neuropsychiatric Genetics and Genomics, Division of Psychological Medicine and Clinical Neurosciences, School of Medicine, Cardiff University, Cardiff, United Kingdom

**Corresponding Author**:

Amy J Lynham

Division of Psychological Medicine and Clinical Neurosciences, School of Medicine

Cardiff University

Hadyn Ellis Building, Maindy Road

Cardiff

CF24 4HQ

### Supplementary Table S1: Potential Covariates

| | HealthWise Wales | | | NCMH | | |
|---|---|---|---|---|---|---|
| **Cognitive Function (‘g’)** | **B** | **95% CI** | **P** | **B** | **95% CI** | **P** |
| HADS | -0.03 | -0.04, -0.03 | <0.001 | -0.05 | -0.06, -0.04 | <0.001 |
| BMI | -0.02 | -0.02-0.006 | 0.005 | | | |
| Alcohol use | 0.13 | -0.0002, 0.26 | 0.05 | 0.51 | 0.26, 0.76 | <0.001 |
| Smoking | -0.1 | -0.2, -0.01 | 0.04 | -0.32 | -0.57, -0.04 | 0.02 |
| Physical activity (ref: inactive) | | | | | | |
| Moderately inactive | 0.14 | -0.03, 0.31 | 0.11 | | | |
| Moderately active | 0.08 | -0.06, 0.23 | 0.27 | | | |
| Active | 0.14 | 0.01, 0.27 | 0.03 | | | |
| Degree | 0.58 | 0.5, 0.66 | <0.001 | 0.84 | 0.58, 1.1 | <0.001 |
| Psychosis | | | | -1.10 | -1.52, -0.68 | <0.001 |
| Bipolar Disorder | | | | -0.46 | -0.78, -0.15 | 0.004 |
| Depression | -0.31 | -0.42, -0.19 | <0.001 | 0.49 | 0.24, 0.74 | <0.001 |
| Anxiety | | | | 0.29 | 0.02, 0.56 | 0.03 |
| OCD | | | | 0.48 | -0.16, 1.13 | 0.14 |
| PTSD | | | | -0.49 | -1.04, 0.05 | 0.07 |
| Eating Disorder | | | | 0.38 | -0.2, 0.96 | 0.2 |
| Personality Disorder | | | | -0.32 | -0.91, 0.26 | 0.28 |
| Alcohol/Substance Dependence | | | | 0.52 | -0.49, 1.52 | 0.31 |
| Autism Spectrum Disorder | | | | 0.35 | -0.37, 1.07 | 0.34 |
| ADHD | | | | 1.26 | 0.16, 2.36 | 0.02 |
| Other Mental Health Problem | | | | 0.06 | -0.37, 0.5 | 0.77 |
| **WHODAS** | **B** | **95% CI** | **P** | **B** | **95% CI** | **P** |
| HADS | 0.05 | 0.05, 0.05 | <0.001 | 0.05 | 0.04, 0.05 | <0.001 |
| BMI | 0.03 | 0.01, 0.04 | <0.001 | | | |
| Alcohol use | -0.08 | -0.2, 0.03 | 0.16 | -0.2 | -0.28, -0.13 | <0.001 |
| Smoking | 0.2 | 0.12, 0.28 | <0.001 | 0.21 | 0.14, 0.29 | <0.001 |
| Physical activity (ref: inactive) | | | | | | |
| Moderately inactive | -0.15 | -0.29, -0.02 | 0.03 | | | |
| Moderately active | -0.15 | -0.27, -0.02 | 0.02 | | | |
| Active | -0.31 | -0.41, -0.2 | <0.001 | | | |
| Degree | -0.26 | -0.33, -0.18 | <0.001 | -0.2 | -0.28, -0.13 | <0.001 |
| Psychosis | | | | 0.05 | -0.11, 0.2 | 0.55 |
| Bipolar Disorder | | | | 0.2 | 0.1, 0.3 | <0.001 |
| Depression | 0.54 | 0.46, 0.63 | <0.001 | 0.04 | -0.03, 0.12 | 0.25 |

| | | | | | | |
|---|---|---|---|---|---|---|
| Anxiety | | | | 0.06 | -0.01, 0.14 | 0.1 |
| OCD | | | | 0.15 | -0.03, 0.32 | 0.1 |
| PTSD | | | | 0.25 | 0.1, 0.4 | 0.001 |
| Eating Disorder | | | | -0.01 | -0.17, 0.15 | 0.91 |
| Personality Disorder | | | | 0.44 | 0.27, 0.61 | <0.001 |
| Alcohol/Substance Dependence | | | | 0.21 | -0.1, 0.53 | 0.19 |
| Autism Spectrum Disorder | | | | 0.41 | 0.22, 0.6 | <0.001 |
| ADHD | | | | 0.004 | -0.28, 0.28 | 0.98 |
| Other Mental Health Problem | | | | 0.04 | -0.08, 0.16 | 0.5 |
| **Employed or in education** | **OR** | **95% CIs** | **P** | **OR** | **95% CI** | **P** |
| HADS | 0.92 | 0.9, 0.95 | <0.001 | 0.92 | 0.9, 0.93 | <0.001 |
| BMI | 0.95 | 0.92, 0.99 | 0.006 | | | |
| Alcohol use | 1.09 | 0.83, 1.45 | 0.52 | 2.05 | 1.51, 2.77 | <0.001 |
| Smoking | 0.7 | 0.56, 0.86 | <0.001 | 0.56 | 0.41, 0.76 | <0.001 |
| Physical activity (ref: inactive) | | | | | | |
| Moderately inactive | 2.59 | 1.78, 3.8 | <0.001 | | | |
| Moderately active | 2.4 | 1.72, 3.36 | <0.001 | | | |
| Active | 3.26 | 2.45, 4.35 | <0.001 | | | |
| Degree | 1.78 | 1.44, 2.22 | <0.001 | 2.82 | 2.06, 3.89 | <0.001 |
| Psychosis | | | | 0.38 | 0.22, 0.68 | 0.001 |
| Bipolar Disorder | | | | 0.42 | 0.29, 0.62 | <0.001 |
| Depression | 0.38 | 0.29, 0.5 | <0.001 | 1.69 | 1.25, 2.28 | <0.001 |
| Anxiety | | | | 1.34 | 0.99, 1.85 | 0.06 |
| OCD | | | | 0.7 | 0.37, 1.36 | 0.29 |
| PTSD | | | | 0.73 | 0.41, 1.29 | 0.27 |
| Eating Disorder | | | | 1.08 | 0.6, 2.02 | 0.8 |
| Personality Disorder | | | | 0.29 | 0.15, 0.53 | <0.001 |
| Alcohol/Substance Dependence | | | | 1.11 | 0.33, 4.33 | 0.87 |
| Autism Spectrum Disorder | | | | 0.8 | 0.4, 1.66 | 0.55 |
| ADHD | | | | 1.69 | 0.58, 6.11 | 0.37 |
| Other Mental Health Problem | | | | 1.19 | 0.73, 1.97 | 0.5 |
| **Living with a partner or spouse** | **OR** | **95% CIs** | **P** | **OR** | **95% CI** | **P** |
| HADS | 0.98 | 0.95, 1 | 0.07 | | | |
| BMI | 1.02 | 0.99, 1.05 | 0.23 | | | |
| Alcohol use | 1.07 | 0.75, 1.55 | 0.72 | | | |
| Smoking | 0.74 | 0.58, 0.94 | 0.01 | | | |
| Physical activity (ref: inactive) | | | | | | |
| Moderately inactive | 1.24 | 0.83, 1.87 | 0.3 | | | |

| | | | |
|---|---|---|---|
| Moderately active | 0.88 | 0.62, 1.24 | 0.47 |
| Active | 1.29 | 0.95, 1.75 | 0.11 |
| Degree | 1.44 | 1.13, 1.85 | 0.004 |
| Anxiety / depression | 0.81 | 0.59, 1.13 | 0.21 |

Shaded cells indicate statistically significant results. Abbreviations from top left to bottom right: NCMH: National Centre for Mental Health; HADS: Hospital Anxiety and Depression Scale; OCD: Obsessive Compulsive Disorder; PTSD: Post-Traumatic Stress Disorder; ADHD: Attention Deficit Hyperactivity Disorder; WHODAS: World Health Organisation Disability Assessment Schedule (12-item). Statistics reported: B: linear regression coefficient; OR: odds ratio; 95% CI: 95% confidence intervals; P: p value (not corrected for multiple testing).

**Supplementary Table S2: Comparison of HealthWise Wales participants who completed or did not complete the WHODAS**

| | WHODAS Completed | WHODAS Not Completed | Single Model | | Model Adjusted for Age, Gender & Device | |
|---|---|---|---|---|---|---|
| **N** | 1033 | 2646 | | | | |
| **Continuous variables** | **Mean (SD)** | **Mean (SD)** | **B** | **P** | **B** | **P** |
| **Age** | 50.97 (16.03) | 57.78 (14.20) | -6.82 | <0.001 | | |
| **Cognitive tasks** | | | | | | |
| Digit Symbol Coding | 0.29 (1.05) | -0.11 (0.96) | 0.4 | <0.001 | 0.08 | 0.01 |
| Backward Digit Span | 0.1 (1.04) | -0.05 (0.98) | 0.15 | <0.001 | 0.07 | 0.12 |
| Vocabulary | -0.1 (1.03) | 0.05 (0.98) | 0.14 | <0.001 | 0.07 | 0.1 |
| Morphed Emotion | 0.17 (1.02) | -0.09 (0.97) | 0.27 | <0.001 | 0.04 | 0.38 |
| Matrix Reasoning | 0.11 (0.99) | -0.07 (1) | 0.18 | <0.001 | 0.05 | 0.26 |
| **BMI** | 26.81 (5) | 26.9 (4.79) | -0.09 | 0.65 | | |
| **Categorical variables** | **N** | **N** | **$X^2$** | **P** | | |
| **Women[1]** | 760 | 1793 | 11.04 | <0.001 | | |
| **Degree** | 567 | 443 | 12.61 | <0.001 | | |
| **Device used** | | | 10.91 | 0.004 | | |
| Laptop / desktop | 519 | 1262 | | | | |
| Tablet | 189 | 614 | | | | |
| Smartphone | 325 | 763 | | | | |
| **Smoking** | 395 | 1120 | 6.01 | 0.01 | | |
| **Alcohol[2]** | 141 | 418 | 2.53 | 0.11 | | |
| **Physical Activity** | | | 1.15 | 0.76 | | |
| Inactive | 194 | 508 | | | | |
| Moderately inactive | 130 | 300 | | | | |
| Moderately active | 196 | 516 | | | | |
| Active | 434 | 1098 | | | | |
| **Depression / Anxiety** | 207 | 423 | 6.43 | 0.01 | | |

All participants from HealthWise Wales who completed at least one cognitive task were included in these comparisons.

### Supplementary Table S3: WHODAS Confirmatory Factor Analysis

| | HealthWise Wales | | | NCMH | | |
|---|---|---|---|---|---|---|
| **Statistic** | **Model 1** | **Model 2** | **Model 3** | **Model 1** | **Model 2** | **Model 3** |
| Comparative fit index (CFI) | 0.981 | 0.992 | 1 | 0.982 | 0.992 | 0.997 |
| Tucker-Lewis index (TLI) | 0.977 | 0.989 | 1.002 | 0.979 | 0.989 | 0.996 |
| Root mean square error of approximation (RMSEA) | 0.055 | 0.038 | <0.001 | 0.075 | 0.053 | 0.033 |
| Standardised root mean square residual (SRMR) | 0.087 | 0.062 | 0.032 | 0.081 | 0.053 | 0.037 |

Results of three confirmatory factor analyses (Diagonally Weighted Least Squares method). Model 1 = One factor model (overall functional impairment), Model 2 = Two-level model (overall functional impairment and WHODAS domains), Model 3 = Two-level model allowing for covariation. Thresholds: CFI>0.9; TLI>0.9; RMSEA<0.08; SRMR<0.08. Shaded cells indicate those that met the required criteria.

### Supplementary Table S4: Associations between individual tasks & measures of functioning

| | HealthWise Wales | | | | | | | | | NCMH | | | | | |
|---|---|---|---|---|---|---|---|---|---|---|---|---|---|---|---|
| | WHODAS | | | Live-in partner or spouse | | | Employed or in education | | | WHODAS | | | Employed or in education | | |
| | B | 95% CI | P | OR | 95% CI | P | OR | 95% CI | P | B | 95% CI | P | OR | 95% CI | P |
| **DSC** | -0.18 | -0.23, -0.13 | <0.001 | 1.29 | 1.09, 1.53 | 0.003 | 1.36 | 1.18, 1.57 | <0.001 | -0.15 | -0.19, -0.11 | <0.001 | 1.78 | 1.48, 2.15 | <0.001 |
| **BDS** | -0.09 | -0.12, -0.05 | <0.001 | 1.17 | 1.03, 1.35 | 0.02 | 1.16 | 1.04, 1.31 | 0.01 | -0.1 | -0.15, -0.06 | <0.001 | 1.4 | 1.17, 1.68 | <0.001 |
| **Vocab** | -0.06 | -0.11, -0.02 | 0.003 | 1.22 | 1.05, 1.41 | 0.01 | 1.02 | 0.89, 1.16 | 0.78 | -0.09 | -0.13, -0.04 | <0.001 | 1.07 | 0.88, 1.29 | 0.5 |
| **MorphEmo** | -0.07 | -0.12, -0.03 | 0.002 | 1.09 | 0.92, 1.27 | 0.32 | 1.24 | 1.08, 1.43 | 0.002 | -0.11 | -0.15, -0.06 | <0.001 | 1.18 | 0.98, 1.43 | 0.08 |
| **Matrix** | -0.12 | -0.16, -0.08 | <0.001 | 1.13 | 0.97, 1.31 | 0.12 | 1.16 | 1.02, 1.32 | 0.02 | -0.1 | -0.14, -0.07 | <0.001 | 1.36 | 1.17, 1.6 | <0.001 |
| **MOT** | | | | | | | | | | -0.12 | -0.16, -0.08 | <0.001 | 1.3 | 1.11, 1.53 | 0.004 |
| **Hartshorne** | | | | | | | | | | -0.11 | -0.16, -0.07 | <0.001 | 1.25 | 1.05, 1.5 | 0.03 |
| **VPAL** | | | | | | | | | | -0.11 | -0.16, -0.07 | <0.001 | 1.41 | 1.17, 1.73 | 0.001 |
| **BART** | | | | | | | | | | -0.07 | -0.11, -0.03 | <0.001 | 1.2 | 1.04, 1.39 | 0.02S |

Shaded cells indicate statistically significant results. Data on live-in partner or spouse was not available for NCMH. Abbreviations: DSC: Digit Symbol Coding; BDS: Backward Digit Span; Vocab: Vocabulary; MorphEmo: Morphed Emotion Identification; Matrix: Matrix Reasoning; MOT: Multiple Object Tracking; Hartshorne: Hartshorne Working Memory Test; VPAL: Verbal Paired Associates Learning; BART: Balloon Analogue Risk Task; WHODAS: World Health Organization Disability Assessment Schedule (12-item); NCMH: National Centre for Mental Health. Statistics reported: B: linear regression coefficient; OR: odds ratio; SE: standard error; 95% CIs: 95% confidence intervals; P: p value (FDR corrected)

**Supplementary Table S5: Associations between overall cognitive function and WHODAS domains (as binary)**

| Domain | HealthWise Wales | | | | | NCMH | | | | |
|---|---|---|---|---|---|---|---|---|---|---|
| | Functional Impairment (n) | No Functional Impairment (n) | OR | 95% CI | P | Functional Impairment (n) | No Functional Impairment (n) | OR | 95% CI | P |
| **All Participants** | | | | | | | | | | |
| **Mobility** | 448 | 586 | 0.77 | 0.7, 0.86 | <0.001 | 591 | 376 | 0.73 | 0.66, 0.81 | <0.001 |
| **Self-care** | 228 | 806 | 0.68 | 0.6, 0.77 | <0.001 | 394 | 572 | 0.82 | 0.75, 0.89 | <0.001 |
| **Household** | 624 | 410 | 0.83 | 0.75, 0.93 | <0.001 | 780 | 187 | 0.78 | 0.69, 0.88 | <0.001 |
| **Cognition** | 598 | 436 | 0.82 | 0.73, 0.91 | <0.001 | 738 | 231 | 0.71 | 0.63, 0.79 | <0.001 |
| **Social** | 489 | 545 | 0.83 | 0.75, 0.92 | <0.001 | 741 | 226 | 0.81 | 0.73, 0.91 | <0.001 |
| **Society** | 694 | 340 | 0.76 | 0.68, 0.85 | <0.001 | 859 | 111 | 0.65 | 0.55, 0.77 | <0.001 |
| **Participants aged 65 years and younger** | | | | | | | | | | |
| **Mobility** | 338 | 464 | 0.75 | 0.67, 0.85 | <0.001 | 533 | 333 | 0.74 | 0.67, 0.82 | <0.001 |
| **Self-care** | 185 | 617 | 0.66 | 0.57, 0.76 | <0.001 | 375 | 490 | 0.83 | 0.76, 0.91 | <0.001 |
| **Household** | 522 | 280 | 0.83 | 0.73, 0.94 | 0.003 | 726 | 142 | 0.81 | 0.71, 0.92 | 0.001 |
| **Cognition** | 507 | 295 | 0.81 | 0.72, 0.92 | 0.001 | 691 | 176 | 0.73 | 0.64, 0.82 | <0.001 |
| **Social** | 420 | 382 | 0.81 | 0.72, 0.92 | <0.001 | 693 | 173 | 0.83 | 0.74, 0.93 | 0.002 |
| **Society** | 557 | 245 | 0.72 | 0.63, 0.81 | <0.001 | 782 | 86 | 0.68 | 0.57, 0.8 | <0.001 |

Each WHODAS domain was dichotomised into any functional impairment versus no functional impairment. Shaded cells indicate statistically significant results. Statistics reported: OR: odds ratio; 95% CI: 95% confidence intervals; P: p values (FDR corrected)

**Supplementary Table S6: Associations between cognition and individual domains of functioning after inclusion of covariates**

| Domain | HealthWise Wales | | | National Centre for Mental Health | | |
|---|---|---|---|---|---|---|
| | B | 95% CI | P | B | 95% CI | P |
| **Cognitive Function 'g'** | | | | | | |
| **Mobility** | -0.1 | -0.18, -0.01 | 0.02 | -0.06 | -0.09, -0.02 | 0.003 |
| **Self-care** | -0.08 | -0.14, -0.01 | 0.02 | -0.05 | -0.08, -0.01 | 0.008 |
| **Household** | -0.06 | -0.11, -0.004 | 0.04 | -0.04 | -0.07, -0.02 | 0.001 |
| **Cognition** | -0.06 | -0.1, -0.02 | 0.007 | -0.05 | -0.08, -0.03 | <0.001 |
| **Social** | -0.05 | -0.11, 0.02 | 0.14 | -0.06 | -0.09, -0.03 | <0.001 |
| **Society** | -0.07 | -0.12, -0.01 | 0.02 | -0.06 | -0.09, -0.03 | <0.001 |

### Supplementary Figure 1: Path diagram of a second-order factor model for the WHODAS

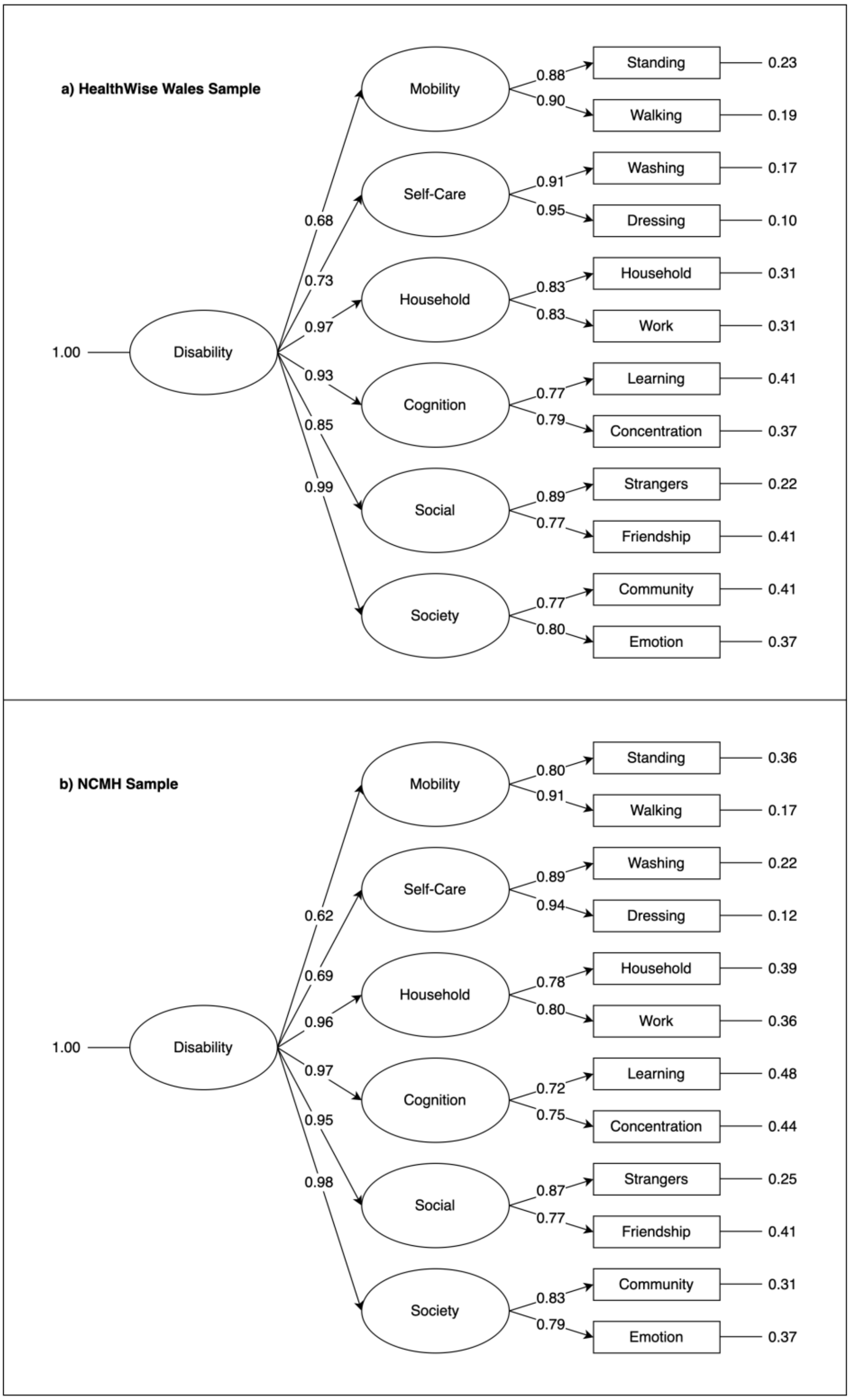

Structure of WHODAS domains derived by confirmatory factor analysis (CFA) in the HealthWise Wales and National Centre for Mental Health (NCMH) samples. Boxes on the right denote the contributing questions from the WHODAS (12 items). Circles represent the latent factors created by the CFA. The arrows represent the standardised factor loadings.

**Supplementary Note 1: NCMH Diagnoses**

As part of CONCA, all participants were asked, "Has a doctor or health professional ever told you that you have any of the following diagnoses?" and given a list of diagnoses to choose from. Participants could select more than one diagnosis. This data was supplemented by diagnosis data that had been collected as part of NCMH's ongoing research to ensure we utilised all available information on participants' historical diagnoses. Data was obtained from the following sources: the NCMH brief assessment, the NCMH Mood and Psychosis Assessment and the NCMH Sleep Study. Information on the different sources of diagnosis within NCMH has previously been published in Woolway et al. (Woolway et al., 2024).

**NCMH Brief Assessment**

This assessment is used when a participant first joins the NCMH cohort. The brief assessment can be completed online or face to face with a researcher. Participants were asked whether a doctor or health professional had ever told the participant that they had a mental health diagnosis and were given a list of psychiatric diagnoses to choose from (see list of diagnoses below). Participants selected all diagnoses that applied.

**NCMH Mood and Psychosis Assessment**

Participants who self-reported a diagnosis of schizophrenia-spectrum disorder, other psychosis or mood disorders were invited to take part in an in-depth assessment. These participants completed a research interview based on the Schedules for Clinical Assessment in Neuropsychiatry (SCAN) (Wing et al., 1990). There were 38 participants in CONCA with available diagnoses from a completed SCAN interview.

**NCMH Sleep Study**

The NCMH sleep study aimed to investigate the relationship between sleep and circadian rhythm disruption and mental health. Participants aged 18 years and older were recruited from the NCMH cohort and completed the Mini-International Neuropsychiatric Interview (MINI) (Sheehan et al., 1998). There were 44 participants in CONCA with available diagnoses from a completed MINI.

**Dealing with data from multiple sources**

We used a hierarchical approach whereby if participants had diagnoses from a semi-structured interview, this data was prioritised over self-report and self-report was only used for unrelated diagnoses. For example, if a participant self-reported psychosis in CONCA or the NCMH brief assessment but their data from the mood and psychosis interview indicated they did not have a psychotic diagnosis then they would not be rated as having psychosis in our analyses. However, they could still be rated as having anxiety if they self-reported a diagnosis of generalised anxiety disorder, as the mood and psychosis interview does not assess symptoms of anxiety.

**Has a doctor or health professional ever told you that you have any of the following diagnoses? (tick all that apply)**

- ☐ Attention Deficit Hyperactivity Disorder (ADHD)
- ☐ Autism
- ☐ Asperger's or other Autism Spectrum Disorder (e.g. Pervasive Developmental Disorder)
- ☐ Dyslexia
- ☐ Dyspraxia
- ☐ Conduct Disorder
- ☐ Oppositional Defiant Disorder (ODD)
- ☐ Tic Disorders
- ☐ Tourette's Disorder
- ☐ Intellectual Disability (ID) / Learning Disability (LD)
- ☐ Depression
- ☐ Bipolar Disorder / Manic Depression
- ☐ Mania / Hypomania
- ☐ Schizoaffective Disorder
- ☐ Psychosis
- ☐ Schizophrenia
- ☐ Postnatal Psychosis / Puerperal Psychosis
- ☐ Postnatal Depression
- ☐ Anorexia
- ☐ Bulimia
- ☐ Obsessive Compulsive Disorder (OCD)
- ☐ Agoraphobia
- ☐ Panic Disorder
- ☐ Phobias
- ☐ Anxiety
- ☐ Borderline Personality Disorder
- ☐ Other Personality Disorder
- ☐ Post-Traumatic Stress Disorder (PTSD)
- ☐ Alzheimer's Disease
- ☐ Other dementia
- ☐ Alcohol Abuse / Misuse
- ☐ Other substance Abuse / Misuse (please specify) ____________
- ☐ Genetic Syndrome (e.g. VCFS) (please specify) ____________
- ☐ Self-harm or suicide attempts
- ☐ Other (please specify) ____________

Sheehan, D. V., Lecrubier, Y., Sheehan, K. H., Amorim, P., Janavs, J., Weiller, E., . . . Dunbar, G. C. (1998). The Mini-International Neuropsychiatric Interview (M.I.N.I.): the development and validation of a structured diagnostic psychiatric interview for DSM-IV and ICD-10. *Journal of Clinical Psychiatry, 59*, 22-33.

Wing, J. K., Babor, T., Brugha, T., Burke, J., Cooper, J., Giel, R., . . . Sartorius, N. (1990). SCAN: Schedules for Clinical Assessment in Neuropsychiatry. *Archives of General Psychiatry, 47*(6), 589-593. Retrieved from http://archpsyc.jamanetwork.com/article.aspx?articleid=495050

Woolway, G. E., Legge, S. E., Lynham, A. J., Smart, S. E., Hubbard, L., Daniel, E. R., . . . Walters, J. T. R. (2024). Assessing the validity of a self-reported clinical diagnosis of schizophrenia. *Schizophrenia, 10*(1), 99. doi:10.1038/s41537-024-00526-5
